# Predicting Cognitive Function Using Transformer-Derived Speech Representations and Longitudinal Coherence Features

**DOI:** 10.64898/2026.08.04.26359730

**Authors:** Deeptanshu Devatha, Joe Xiao

## Abstract

Dementia affects more than 55 million people worldwide, and its progressive decline is difficult to track using infrequent in-person assessments, which can miss subtle changes between visits and add to clinician burden. One widely used measure, the Mini-Mental State Examination (MMSE), is administered intermittently and remains subject to inconsistent scoring judgment, limiting early detection. Prior speech-based machine learning approaches have largely focused on cross-sectional classification rather than longitudinal cognitive forecasting. We introduce a longitudinal, patient-level framework that combines transformer-derived semantic speech representations with longitudinal speech-change features and clinical history to forecast a patient’s future MMSE score from their history of prior visits. To our knowledge, this is the first framework to unite transformer-derived speech encoding with longitudinal forecasting of cognitive severity, rather than single-visit classification alone. We evaluate this framework on longitudinal transcripts from the DementiaBank Pitt Corpus using a LightGBM gradient-boosted regression model, validated with patient-grouped cross-validation to prevent identity leakage between training and evaluation folds. The model forecasts future MMSE scores with high accuracy and stability across folds (*R*^2^ = 0.840 *±* 0.015, *RMSE* = 2.75, *MAE* = 2.02, Pearson *r* = 0.917), with transformer-derived speech representations contributing meaningful predictive signal alongside clinical history. Ablation analysis demonstrated that transformer-derived speech representations provided complementary predictive information beyond clinical variables. These results establish speech as a viable longitudinal digital biomarker of cognitive decline, offering a low-burden complement to intermittent clinical assessment that could enable earlier detection and more frequent monitoring, supporting better-timed care decisions for patients with dementia.

## 1 Introduction

Dementia is a rising global problem that affects over 55 million people worldwide (World Health Organization 2023), with around 7.4 million Americans over the age of 65 living with the condition (Alzheimer’s Association 2026). Current non-computational diagnostic methods for detecting dementia severity, such as in-person clinical exams, present notable limitations for tracking cognitive decline. For example, the Mini-Mental State Exam (MMSE), one of the most widely used tests of cognitive function (Folstein et al. 1975), is usually administered every 6 months. Because cognitive tests as such are not given very frequently, clinicians have a very limited understanding of dementia patients’ cognitive function; subtle warning signs of cognitive decline and minor changes in brain condition may not be captured. Sparsely conducted tests make early detection nearly impossible. To continue, because clinical cognitive function exams are administered by a human clinician, other confounding variables, such as human biases, pose problems, such as inconsistent judgment when scoring ambiguous or partial-credit responses (Molloy et al. 1991). For example, clinicians may score inconsistently on items such as spelling “world” backwards or sentence construction. Additionally, clinician workload could increase fatigue during test administration and, consequently, raise the risk of error, such as skipped test items, mistimed recall intervals, or scoring mistakes (Machizawa et al. 2025).

Using speech as a biomarker of cognitive decline across various time points, combined with machine-learning analysis, addresses these problems and provides a reliable method for predicting cognitive function in patients with dementia. Machine learning analysis of dementia patients’ speech may enable more frequent assessment than administering full cognitive tests, since speech samples can potentially be collected and analyzed more quickly, increasing the chance of early prediction of future cognitive decline. Subtle changes in brain function may be important indicators of future cognitive function, and frequent analysis of speech in dementia patients may support more timely monitoring of cognitive change. Additionally, this method of predicting cognitive function could reduce reliance on subjective human scoring judgment, complementing rather than replacing clinician assessment.

Several studies have explored the use of deep learning models to encode dementia patients’ speech and use the encodings to determine cognitive impairments. For example, Yang et al. (2022) conducted a literature review of deep learning approaches for Alzheimer’s disease detection using patient speech, finding that deep learning methods consistently outperformed machine learning approaches and that transformer models generally performed the best in Alzheimer’s classification. To continue, Qiao et al. (2021) explored the combination of handcrafted linguistic features derived from spontaneous patient speech with pre-trained transformer language models for detecting Alzheimer’s disease. They found that transformer embeddings, combined with handcrafted linguistic features, increased prediction accuracy, and that this hybrid framework outperformed models relying solely on handcrafted features. Similarly, Balagopalan et al. (2021) compared pre-trained language foundation models, rather than analysis using handcrafted linguistic features, given the task of Alzheimer’s detection and found strong predictive performance. Additionally, Balagopalan et al. (2020) used various transformer model architectures, such as BERT (Devlin et al. 2019), for dementia detection; compared with traditional NLP techniques for the same task, such as bag-of-words vectorization, transformer model embeddings were more accurate predictors. These studies rely on deep learning models to analyze patient speech and detect dementia. However, most of these studies are cross-sectional and focus on dementia classification rather than on predicting future cognitive decline. In addition, limited research has examined how transformer-derived semantic coherence features change over time across repeated patient visits; most studies focus on single visits rather than temporal progression.

A related area of research uses longitudinal analysis of dementia patient speech to track disease progression over time. Firstly, in a study by Forbes-McKay et al. (2013), spontaneous speech from patients with mild-to-moderate Alzheimer’s disease was assessed at baseline, 6 months, and 12 months, and the prevalence of phonological errors and reduced syntactic complexity increased progressively over the study period. This study demonstrated that speech biomarkers exhibit measurable longitudinal changes across repeated patient visits, suggesting that repeated observations may provide additional information about disease progression beyond a single assessment. Additionally, in a study by Gkoumas et al. (2024), which created a longitudinal multi-modal dataset of speech and text data from dementia patients collected across up to 28 repeated sessions per participant, a similar finding was observed. These longitudinal studies primarily rely on handcrafted linguistic and acoustic features rather than transformer-derived contextual representations. Moreover, most of these studies focus on monitoring disease progression rather than predicting future severity of cognitive decline. These gaps limit the potential of using speech as a biomarker for predicting longitudinal cognitive decline, as transformer models have been shown to achieve higher accuracy in analyses of dementia patients’ speech, suggesting a promising direction for future work.

This study aims to develop a longitudinal machine learning framework to predict future Mini-Mental State Examination (MMSE) scores – our operational measure of cognitive severity – using transformer-derived speech representations, handcrafted discourse features, and decision-tree algorithms. Our primary contribution, to the best of our knowledge, is this longitudinal MMSE-forecasting task itself, rather than the transformer-based speech encoding, which follows established methods; combining established speech representations with longitudinal forecasting, rather than single-visit classification, integrates deep and machine learning with temporal data and handcrafted features for regression analysis.

## 2 Methods

This study used transcripts of speech recordings from the DementiaBank Pitt Corpus (Becker et al. 1994). These recordings were interviews conducted with study participants. The data contain 104 elderly participants serving as controls, 208 participants with dementia, and 85 with an unknown diagnosis (Becker et al. 1994). Data were collected longitudinally; each patient was assessed by an interviewer once a year. Participants whose speech samples were unavailable in the dataset were excluded, and participants with only one visit sample were also excluded to enable longitudinal modeling across time points. Additionally, some samples were excluded because they had missing MMSE scores for their patients’ last visits. As a result, the final training dataset consists of 126 patients: 69 from the Dementia cohort and 57 from the Control cohort, and is skewed towards patients with dementia rather than controls. This is because control participants were typically assessed only once or twice and were more likely to be excluded from the final dataset.

Of these 126 patients, most had 2 or 3 visits, while a small number had 4 or 5 visits (Figure 2). Longitudinal samples were constructed using 1 to 4 prior visits to predict the MMSE at a future visit, yielding 173 prediction samples (Figure 3). Target MMSE scores ranged from 1 to 30 (mean = 23.96, SD = 6.84), with 38.7% of samples falling below the clinical impairment threshold of 24 (Figure 1). Most prediction sequences consisted of a single prior visit. Additional dataset characteristics, such as MMSE score distributions and the correlations between target visit numbers and MMSE scores, are shown in Figures 1–4.

**Fig. 1.**
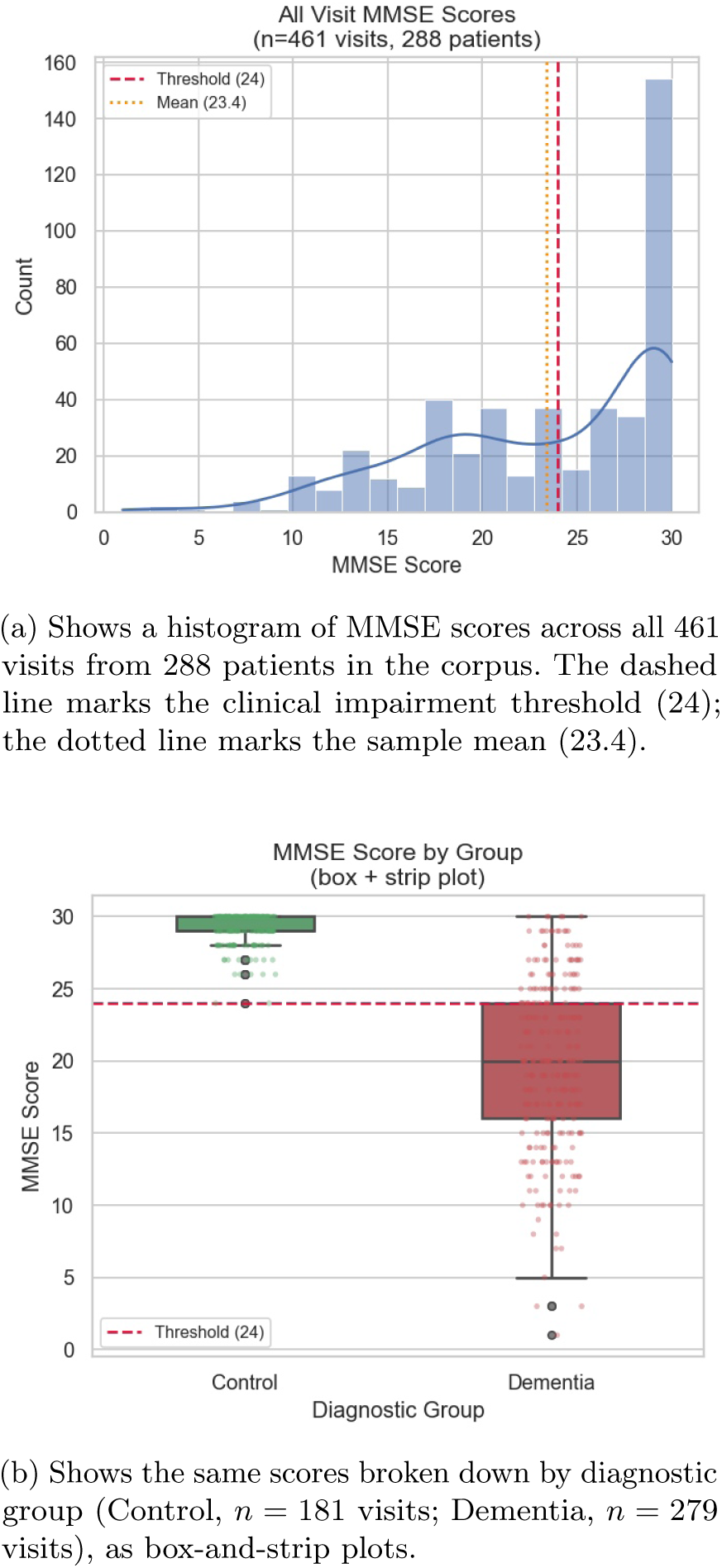
Shows MMSE scores across the full visit-level corpus, split by diagnostic group (a, b). Control-group scores sit tightly near the top of the scale, while Dementia-group scores are lower on average and far more spread out, extending down into severe impairment.

**Fig. 2.**
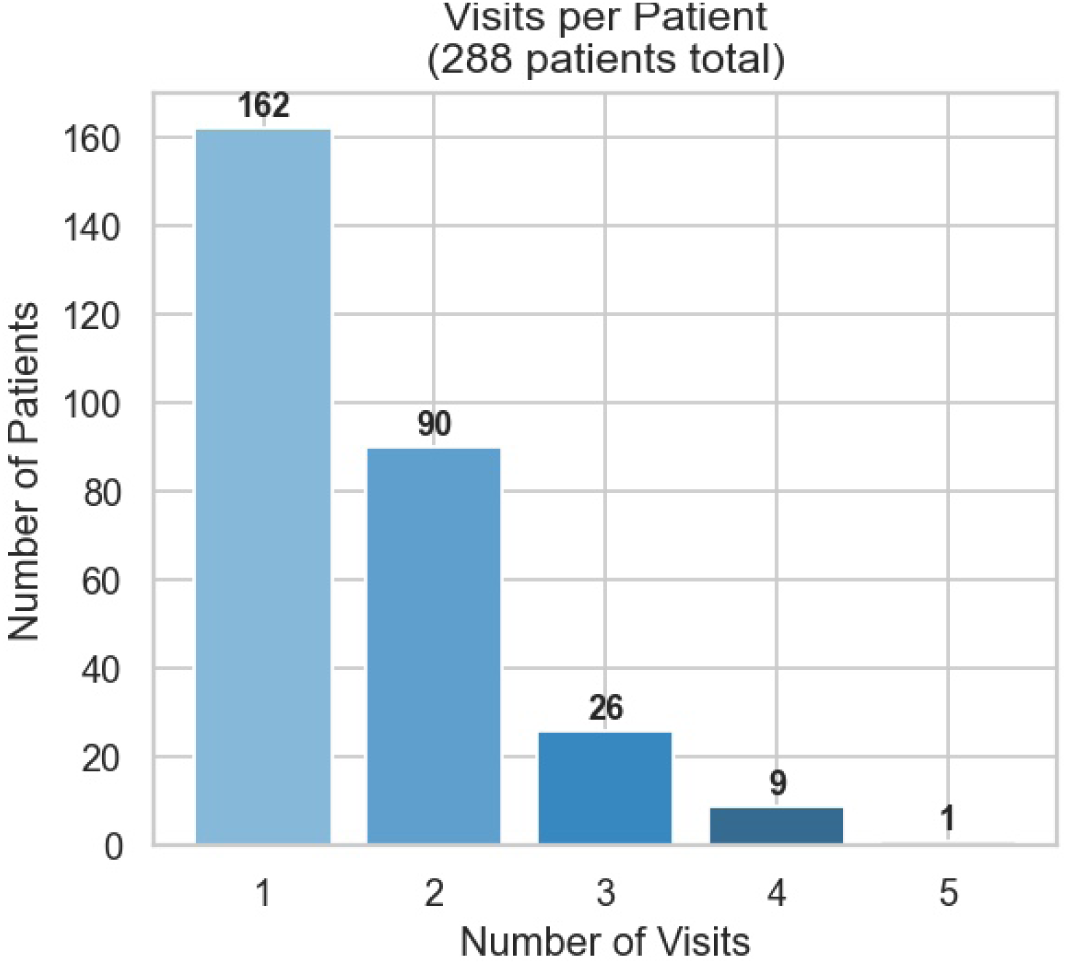
Shows, for each of the 288 patients in the working corpus, the number of visits completed, from one to five. Most patients (162 of 288) were seen only once, leaving a much smaller group with the repeat visits needed for longitudinal modeling.

**Fig. 3.**
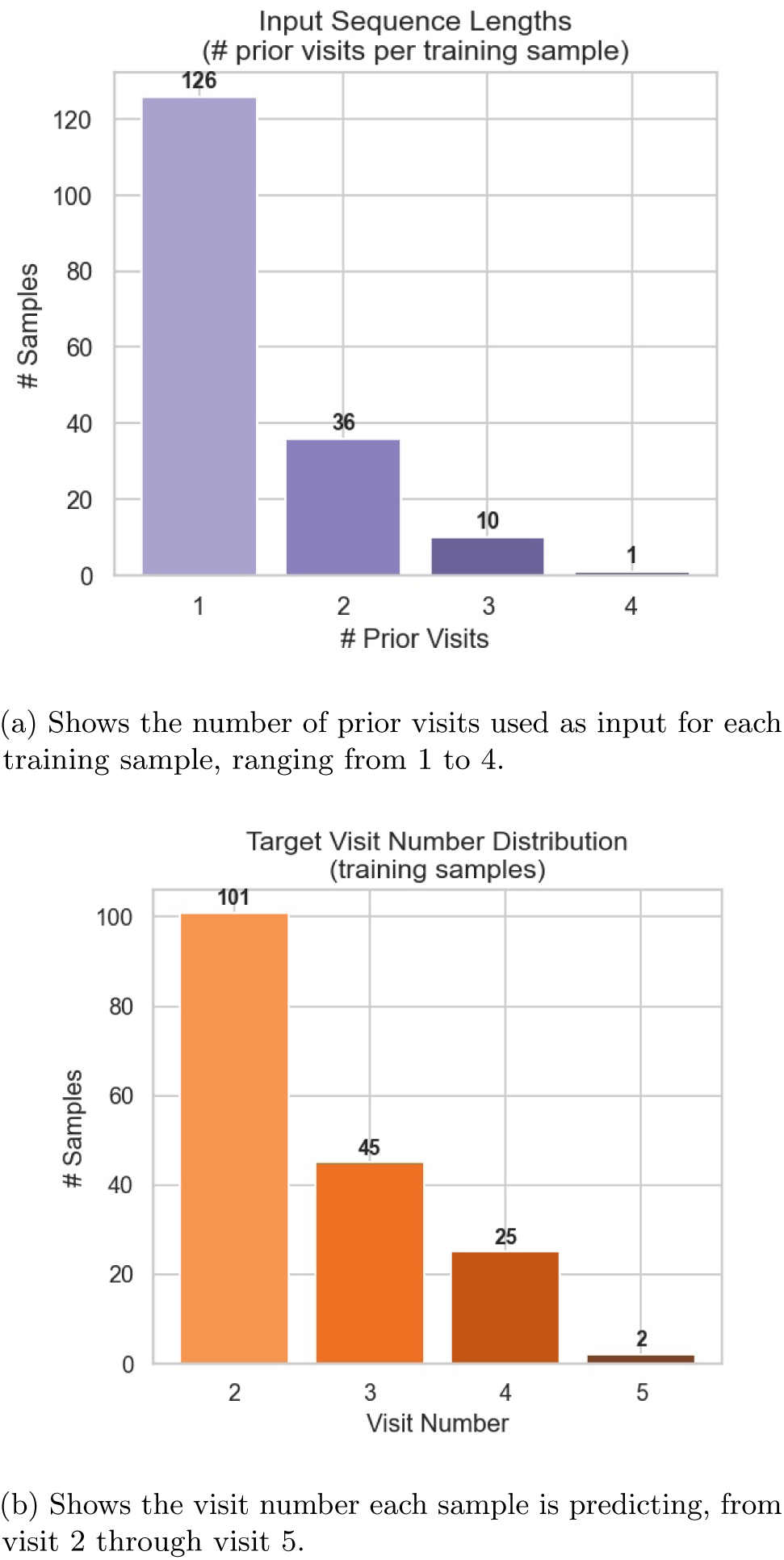
Shows how the 173 training samples were built from patients’ visit histories: how many prior visits fed into each one (a), and which visit it was predicting (b). Most samples use just one prior visit to predict the next one (visit 2), reflecting how short most patients’ visit histories are.

**Fig. 4.**
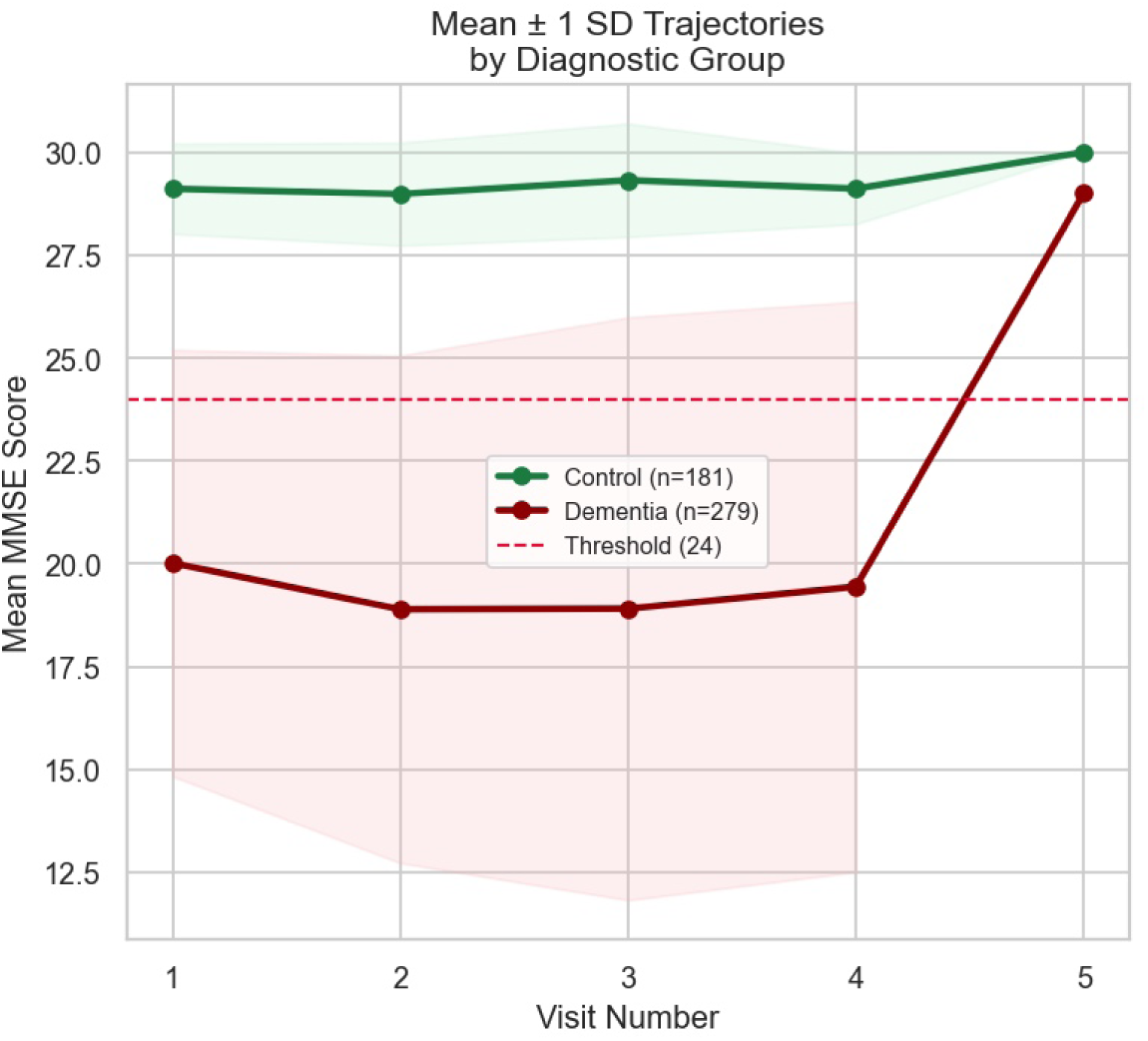
Shows how mean MMSE score changes across visits (1–5) for the Control (*n* = 181 visits) and Dementia (*n* = 279 visits) groups, with shaded bands indicating *±*1 SD and a dashed line marking the clinical impairment threshold (24). Control scores stay flat near the top of the scale across visits, while Dementia scores are consistently lower and more variable, dipping below the impairment threshold at several visits.

### 2.1 Preprocessing

Our next step following data collection was data preprocessing. Firstly, we removed all interviewer speech from the transcripts to ensure that transformer encodings of the transcripts, as well as derived handcrafted features, represent patient speech only. All metadata and timestamps were additionally removed. Next, we performed a token-filtering process that cleaned patient transcripts by retaining only alphabetic words, contractions, and filler tokens (e.g., &-, indicating stutter).

### 2.2 Feature Extraction

Following data preprocessing, we extracted features from the cleaned data. The following features were handcrafted by us and derived from all transcripts in this dataset.

1. Let a transcript contain sentences with SBERT (Reimers and Gurevych 2019) embeddings *S*_1_*, S*_2_*, . . . , S_n_*, with SBERT embeddings *e*_1_*, e*_2_*, . . . , e_n_*. The first feature we decided to extract from the transcripts is Adjacent Cosine Similarity (ACS). This metric measures the congruence between each subsequence of length 2 in the patient transcripts, using cosine similarity.

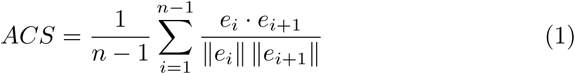

Measuring similarity between adjacent sentences measures topic continuity; i.e., the extent to which a patient can maintain certain topics in speech without abruptly introducing unrelated ones. Higher *ACS* values indicate greater levels of topic continuity in patient speech, representing more coherent speech. We used this metric to measure cognitive decline, as topic maintenance is a major differentiator between normal speech and speech from dementia patients.

2. **Global Coherence (GC).** Along with similarity derived between adjacent sentences, we also included a metric that measures the similarity between the very first and very last sentence of a transcript.

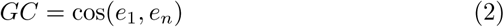

Lower *GC* values indicate topic wandering over the course of the transcript, while higher values indicate that the speaker returned to, or never departed from, the transcript’s opening topic. Where *ACS* captures only local, sentenceto-sentence continuity, *GC* captures whether coherence is maintained across the entire transcript.

3. **Semantic Slope (SS).** Semantic slope captures the trend of semantic coherence as speech progresses within a transcript. Each sentence’s cosine similarity to the transcript’s first sentence, cos(*e*_1_*, e_i_*) for *i* = 2*, . . . , n*, is regressed linearly against that sentence’s position *i* in the text.

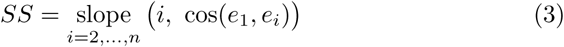

Negative slope values indicate progressive semantic drift away from the transcript’s opening topic, potentially indicating a loss of coherence as the transcript continues.

4. **Topic Drift (TD).** Topic drift quantifies the extent to which a speaker’s individual sentences depart from the transcript’s overall central topic, defined relative to the transcript’s mean embedding 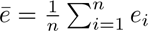.

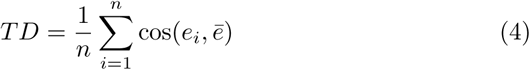

Lower *TD* values indicate that sentences depart, on average, further from the transcript’s central topic — i.e., greater topic drift — whereas higher values indicate stronger maintenance of a central topic throughout the transcript.

5. **Embedding Variance (EV).** Embedding variance, a feature related to Adjacent Cosine Similarity, measures the variance (rather than the mean) of consecutive-sentence similarity across a transcript.

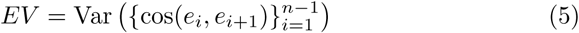

Higher embedding variance values indicate more erratic semantic movement between sentences, showing difficulty in maintaining a single coherent topic. Lower values indicate consistent semantic representation throughout the patient transcript speech.

6. Content-word repetition was also applied in our study as an indicator of topic maintenance. As each token in the transcript is scanned in order, a token is counted as a repeat if that same word has already occurred earlier in the transcript. Let *N* be the total number of tokens in the transcript.

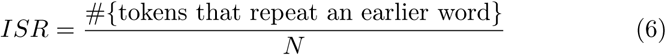

We refer to this metric as Inter-Sentence Repetition (ISR); higher *ISR* values indicate a greater degree of lexical perseveration across the transcript, rather than the introduction of new words and ideas.

7. A frequent pattern in people with cognitive impairment is the use of filler words, such as “um” or “uh”. This indicates disfluency in thinking and speech formation. The metric we use to capture this pattern is the ratio of filler/disfluency tokens to total tokens in the entire transcript. Let *N* be the total number of tokens in a given patient transcript. Let *F* be the total number of filler/disfluency tokens in the transcript. The filler rate *FR* is then defined as:

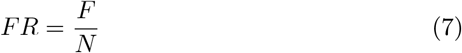

Higher *FR* values indicate a larger use of filler words relative to total words in a patient’s sequence of transcript sentences, representing a greater degree of disfluency in patient speech.

8. **MATTR (Moving-Average Type-Token Ratio).** Beyond filler words, we also measured lexical diversity directly. For a token sequence of length *N ≥* 50 and window size *w* = 50, MATTR is the type-token ratio averaged over all sliding windows of length *w* in the transcript (for *N <* 50, MATTR reduces to the ordinary type-token ratio, |unique tokens*|/N* ).

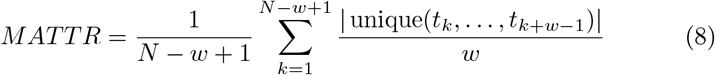

Lower MATTR values indicate less varied vocabulary use, a pattern associated with cognitive decline; the moving-average formulation makes MATTR more robust to differences in transcript length than the ordinary type-token ratio.

9. **Mean Sentence Length (MSL).** As a measure of syntactic complexity, we also computed the mean number of word tokens per utterance across a transcript:

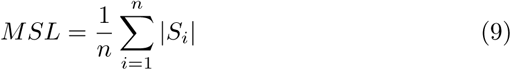

Shorter mean sentence lengths are associated with reduced syntactic complexity in patient speech.

10. **Fragment Ratio (FRAG).** Using spaCy dependency parsing to identify sentence boundaries and part-of-speech tags, we computed the proportion of a transcript’s sentences that contain no verb token, indexing syntactically incomplete utterances:

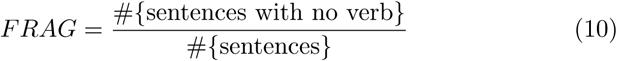

Higher *FRAG* values indicate a greater proportion of syntactically incomplete utterances in patient speech.

11. **Named Entity Rate (NER).** We additionally computed the rate of spaCy- recognized named-entity mentions, *E*, per token:

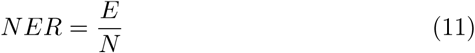

Lower *NER* values indicate reduced use of specific, identifiable entities (people, places, etc.) in speech, another pattern associated with cognitive decline.

12. **Generic Word Rate (GWR).** Finally, we computed the rate of semantically vague, generic words (*thing*, *stuff*, *something*, *that*, *this*) per token:

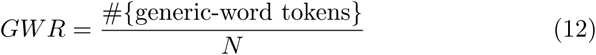

Higher *GWR* values indicate greater reliance on vague, non-specific vocabulary in place of more specific words, a pattern also associated with cognitive decline.

Together with the previously described handcrafted measures, twelve visit-level speech features are used in the prediction of cognitive decline across patient visits, summarized in Table 1.

**Table 1.**
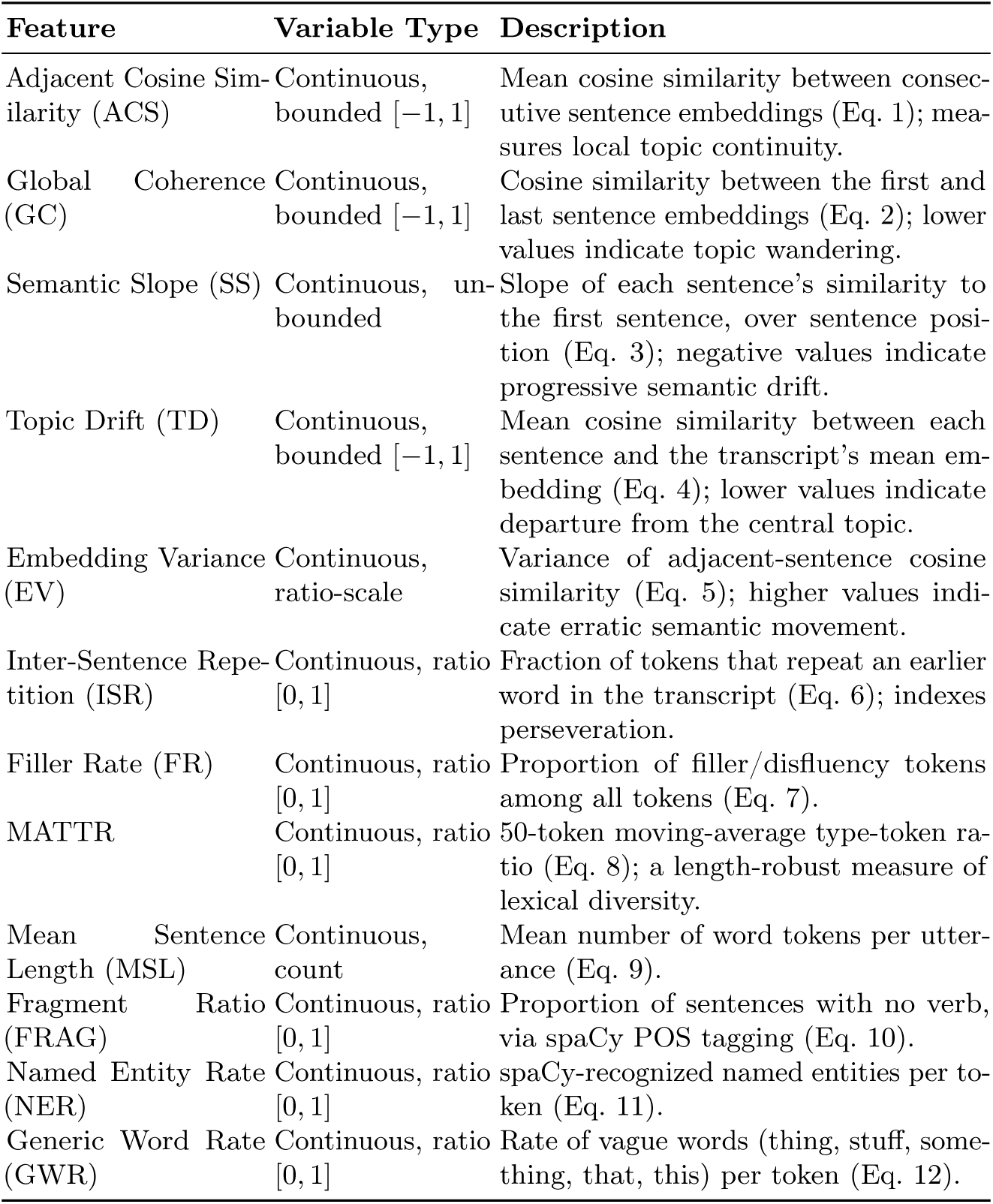
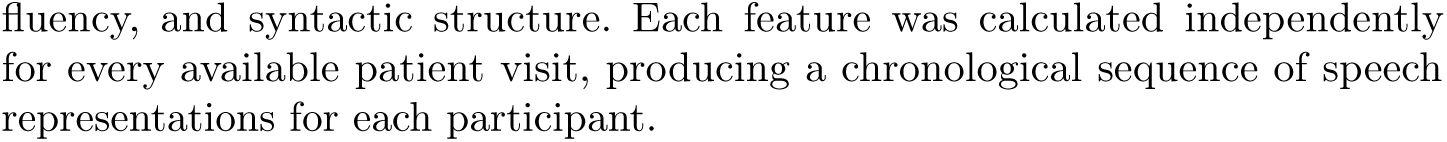
Handcrafted Linguistic Features Extracted per Visit.

### 2.3 Feature Engineering

To create a fixed-dimensional representation of patient visit history for a machine learning prediction task, we summarized each patient’s visit history using recent-state, short-term change, and temporal observation features. First, the 12 linguistic features were extracted from the final visit before MMSE prediction. Importantly, rather than an average across previous visits, these values represent only the final available prior visit, capturing the patient’s current speech state. To capture temporal information, recent two-visit history features were extracted for eight of these twelve speech features: Embedding Variance, Global Coherence, ACS, Topic Drift, MATTR, Mean Sentence Length, Generic Word Rate, and Inter-Sentence Repetition. For each of these eight features *x*, the short-term change *Δx* = *x_t_ − x_t−_*_1_ was computed, where *x_t_* represents the feature value from the most recent visit immediately before the target visit, and *x_t−_*_1_ represents the feature value from the visit immediately preceding the visit represented by *x_t_*. These temporal representations quantify the most recent change in patient speech characteristics, rather than summarizing the entire patient history.

In addition, four metadata variables were included: the number of prior visits available for prediction, the time interval between the target visit and the immediately prior visit, the average time gap between visits, and the total patient observation span. The purpose of these metadata variables is to capture more information about the patient’s observation structure.

Together, the longitudinal speech features, combined with observation-history representations, comprised 24 dimensions: 12 recent linguistic features, 8 short-term linguistic changes, and 4 temporal metadata features.

The second component of the feature representation for this longitudinal prediction task is transformer-derived semantic embeddings extracted from patient speech. Sentence-BERT (SBERT) embeddings (Reimers and Gurevych 2019) were extracted from each patient transcript to capture a context-rich vector representation of speech beyond explicit handcrafted linguistic features. For each prediction sample, past visits were aggregated into a single embedding to represent the patient’s longitudinal speech characteristics. The original aggregated semantic embedding per transcript contained 384 dimensions; to reduce computational cost and because many dimensions contained correlated information, principal component analysis (PCA) was applied to the raw embeddings using the scikit-learn implementation (Pedregosa et al. 2011). The first 20 principal components were retained, yielding a 20-dimensional neural semantic representation of patient speech aggregated across the patient’s visit history.

The final aspect of our feature representation for this task includes a variety of baseline clinical and demographic information for each patient. This information includes each patient’s entry MMSE score (Folstein et al. 1975), their Blessed Dementia Rating score (Blessed et al. 1968), Clinical Dementia Rating (CDR; Hughes et al. 1982), NYU staging score (Reisberg et al. 1982), Mattis Dementia Rating Scale score (Mattis 1988), baseline diagnosis code, age, education, and sex. These 9 features provide context on patients’ baseline cognitive states and certain demographic factors that may impact cognitive function.

All of these feature representation components were represented in the final feature vector, which was formed by horizontally concatenating these three representation blocks. The final vector for each prediction sample comprises 53 dimensions: 12 recent linguistic features, 8 short-term linguistic changes, 4 temporal metadata features, 20 SBERT-PCA components, and 9 baseline clinical and demographic features.

Each row of the resulting training samples matrix represents a prediction task that uses speech-derived measurements, recent changes in patient speech, dense semantic representations, temporal observation characteristics, and baseline clinical information to predict a patient’s MMSE score for the target visit.

### 2.4 Model

These training samples were used as inputs to a supervised machine learning framework trained for the regression task of predicting a given patient’s MMSE score at the target visit. The resulting feature space after our feature construction contains distinct predictors of cognitive decline that may exhibit non-linear relationships; for this reason, a gradient-boosted decision tree regression model was used. The specific tree model is LightGBM (Ke et al. 2017), selected to capture complex relationships across distinct features, model potentially non-linear relationships, and address the small dataset size. This model takes the previously discussed feature vector as input and outputs a continuous prediction of the MMSE score for a given patient.

For model evaluation, 5-fold GroupKFold cross-validation (CV; Pedregosa et al. 2011) was used. During this process, training samples were partitioned by patient ID. In this way, all samples from the same participant were assigned strictly to either the training or validation fold in each CV iteration. This prevents information leakage by ensuring that the model is not trained and validated on data from the same patient, allowing model generalization to unseen patients.

### 2.5 Training Procedure

Before model training, RobustScaler normalization (Pedregosa et al. 2011) was applied to reduce the influence of outliers and account for the heterogeneous feature distribution. Following this, Pearson correlation-based pruning with a correlation threshold of 0.8 was performed to remove redundant predictors. Clinical and demographic variables were retained during this process. After this filtering process, we used a preliminary LightGBM model to perform permutation-importance-based feature selection (Breiman 2001), retaining the 15 most influential features. Clinical and demographic variables excluded from this feature selection process were added back because baseline cognitive and functional measures have been shown to be important predictors of subsequent cognitive trajectories (Xie et al. 2012). Each feature in the resulting feature set was then discretized using KBinsDiscretizer (Pedregosa et al. 2011); continuous values were placed into quantile-based bins. These discrete features were appended alongside the continuous feature values, doubling the dimensions of the final feature matrix. Due to the limited dataset size, Gaussian augmentation was applied to this matrix to expand it with perturbed copies. For each sample, three noised copies were generated. Noise was drawn independently per feature from a zero-mean Gaussian scaled to 2% of that feature’s marginal standard deviation. As a result, the sample pool increased to 4 times its original size. Each synthetic copy was tagged with its parent patient’s ID, and GroupKFold splitting was performed on these tags rather than sample indices, ensuring that all copies of a given patient’s data fell within a single fold and preventing the same patient’s data from leaking across the training and validation splits. Consequently, validation folds during cross-validation contained both original and augmented copies of held-out patients; however, the metrics reported in Table 2 are computed from out-of-fold predictions for the original, unaugmented samples only.

**Table 2.** Model Performance Metrics Across 5-Fold Cross-Validation.

| Metric | Per-fold (mean ± SD) |
| --- | --- |
| $R^2$ | $0.840 \pm 0.015$ |
| $RMSE$ | 2.75 |
| $MAE$ | 2.02 |
| Pearson $r$ | 0.917 |

The hyperparameters of the LightGBM model were optimized using Optuna (Akiba et al. 2019). We used a Tree-structured Parzen Estimator sampler (Bergstra et al. 2011) over 200 trials. During these trials, ten parameters were searched: number of estimators, maximum depth, learning rate, number of leaves, minimum child samples, L1 and L2 regularization coefficients, subsample ratio, feature subsample ratio, and minimum split gain. Each hyperparameter configuration was scored based on the mean validation *R*^2^ score across the 5-fold GroupKFold split.

Following the 200 Optuna trials, the best-performing configuration was used to train 5 LightGBM models with identical hyperparameters and random seed. Each model was trained only on its corresponding fold’s training and validation partitions. The five training partitions pairwise overlap on 3 of 5 patient groups, so the models differ solely in the rotating subset of excluded patients.

The correlation-based pruning, permutation-importance feature selection, PCA dimensionality reduction, and Optuna hyperparameter search described above were each performed once on the full dataset rather than being independently refit within nested inner folds. Because feature selection and hyperparameter tuning in particular are label-dependent, this may introduce a modest optimistic bias in the reported cross-validation metrics (Table 2).

## 3 Results

Model performance metrics were computed independently within each of the 5 cross-validation folds, and summarized as mean *±* standard deviation (Table 2). We used the following metrics to assess model performance: the coefficient of determination (*R*^2^), root mean squared error (*RMSE*), mean absolute error (*MAE*), and Pearson correlation (*r*) between predicted and observed MMSE scores.

The narrow standard deviation observed confirms stable model performance across patient subsets. In sum, this machine learning framework evaluates whether a LightGBM model can use longitudinal speech patterns, semantic representations, temporal context, and clinical information to predict future cognitive function in new patients not observed during training.

A larger proportion of model error occurs for samples with severe impairment (i.e., MMSE *<* 10), due to the smaller training set in this area. Prediction residuals were approximately centered (mean = *−*0.13, SD = 3.35) with a mild negative skew. This indicates a slight model overprediction for severely impaired patients.

### 3.1 Feature Importance

Following model evaluation, feature importance analysis was conducted to determine which features had a greater predictive weight for cognitive decline. Feature importance values were computed across the 5 CV folds and averaged. We used the following per-fold permutation importance procedure (Breiman 2001) to calculate these values:

For a fold *i* with a validation set *V_i_* and a trained model *f_i_*, we first computed the baseline validation *R*^2^ using the following formula:

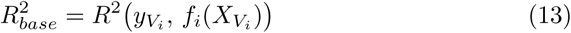

where *y_Vi_* represents the set of output labels for a given fold’s validation set and *X_Vi_* represents the set of features for a given fold’s validation set. To continue, for each feature *j*, we created an altered dataset 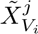 by randomly shuffling column *j* to break that feature’s correlation with the target MMSE score. Then, we computed 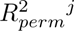, which represents a new *R*^2^ value on the permuted dataset:

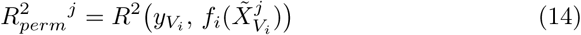

The permutation importance value for a given feature *j* was then computed as the difference between 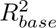 and 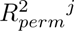, as such:

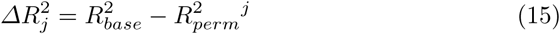

This process was repeated 10 times, and the average value across the 10 trials was taken to reduce stochastic noise from the feature-shuffling step. Finally, to aggregate the feature importance values across CV folds, we

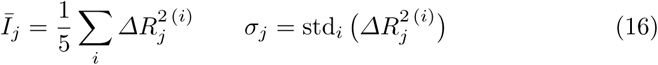

The reported value *̄I_j_* represents the mean drop in *R*^2^ when feature *j*’s signal is destroyed, averaged across all 5 folds. Higher values of *̄I_j_* indicate a greater importance for feature *j*. Because of the noise-reduction step, *̄I_j_* reliably depicts the expected feature importance across different patient subsets in the cohort. Next, the reported *±* standard deviation (*σ_j_*) measures fold-to-fold variability. Features with high *σ* have fold-dependent importance, meaning their relevance depends on which patients are included in the training set.

### 3.2 Ablation Study

To directly test the contribution of each feature source, we ran an ablation study in which a fixed, untuned LightGBM regressor (300 estimators, max depth 3, learning rate 0.05) was trained on isolated feature subsets under the same patient-level 5-fold GroupKFold cross-validation protocol described in Section 2.4, with all samples from the same participant restricted to either the training or validation fold in each iteration.

This ablation design was used to isolate the contribution of each feature source from the impact of hyperparameter tuning and correlation-based pruning used in the main pipeline (Section 2.5); therefore, the “All features combined” row in Table 3 is not meant to be compared directly with the results of the main model pipeline shown in Table 2.

**Table 3.**
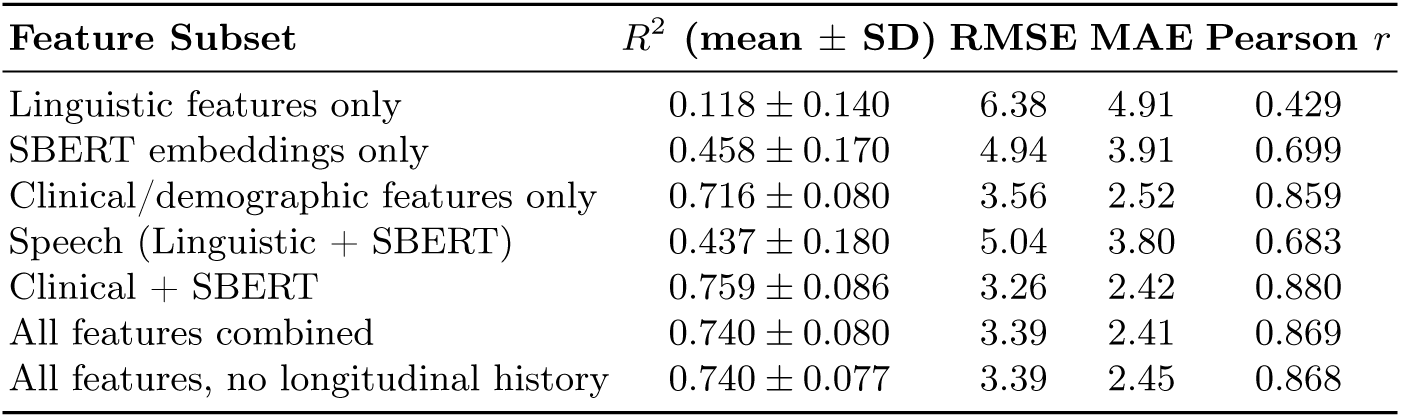
Feature-Source Ablation (fixed, untuned LightGBM; 5-fold GroupKFold C<u>V)</u>.

| Feature Subset | $R^2$ (mean ± SD) | RMSE | MAE | Pearson $r$ |
| --- | --- | --- | --- | --- |
| Linguistic features only | $0.118 \pm 0.140$ | 6.38 | 4.91 | 0.429 |
| SBERT embeddings only | $0.458 \pm 0.170$ | 4.94 | 3.91 | 0.699 |
| Clinical/demographic features only | $0.716 \pm 0.080$ | 3.56 | 2.52 | 0.859 |
| Speech (Linguistic + SBERT) | $0.437 \pm 0.180$ | 5.04 | 3.80 | 0.683 |
| Clinical + SBERT | $0.759 \pm 0.086$ | 3.26 | 2.42 | 0.880 |
| All features combined | $0.740 \pm 0.080$ | 3.39 | 2.41 | 0.869 |
| All features, no longitudinal history | $0.740 \pm 0.077$ | 3.39 | 2.45 | 0.868 |

Clinical and demographic features alone were the strongest single source (*R*^2^ = 0.716), consistent with the dominant permutation importance of entry MMSE and the Blessed score (Section 3.1); permutation importance and this ablation offer complementary evidence, as the former identifies which features are influential within the final trained model while the latter evaluates how much signal each feature group carries when isolated. SBERT embeddings alone (*R*^2^ = 0.458) captured richer semantic information from speech than the handcrafted linguistic features alone (*R*^2^ = 0.118), though neither speech-only source matched clinical features in isolation, and combining the two speech sources did not improve on SBERT alone (*R*^2^ = 0.437 vs. 0.458), suggesting some overlap between them under this fixed-capacity model. Adding SBERT embeddings to clinical features improved performance beyond clinical features alone (*R*^2^ = 0.759), the best result among the feature subsets tested, suggesting that speech representations contribute predictive information that clinical variables alone may not capture. These ablation comparisons reflect point estimates from 5-fold cross-validation rather than formal significance testing, and because the reported standard deviations overlap (0.716 *±* 0.080 vs. 0.759 *±* 0.086), this difference should be read as directional evidence rather than a statistically confirmed effect. The full feature set scored slightly lower (*R*^2^ = 0.740) than Clinical + SBERT; because this ablation uses a fixed, untuned model without the feature selection or hyperparameter tuning applied in the main pipeline, the fixed-capacity model may simply have more difficulty extracting incremental signal from the added linguistic and longitudinal dimensions, rather than these features being unhelpful in the tuned pipeline.

Separately, we tested whether the explicitly engineered longitudinal change features contribute beyond the last-visit snapshot: removing them left performance unchanged (*R*^2^ = 0.740 in both cases). Given the small sample size and the difficulty of isolating sparse, visit-to-visit change with manually engineered delta features, this controlled ablation may simply lack the power to detect their marginal contribution, and should not be read as evidence that longitudinal modeling is unnecessary. Together, these results support speech-derived representations as a complementary, rather than substitute, source of predictive signal alongside clinical history; the higher performance of the main pipeline (*R*^2^ = 0.840; Table 2) reflects the added benefit of hyperparameter optimization and feature selection in integrating these heterogeneous sources beyond what this fixed-capacity ablation setup can achieve.

## 4 Discussion

### 4.1 Summary of Findings

During this study, we demonstrated that longitudinal speech-derived representations can effectively predict future cognitive function, with LightGBM predictions showing a strong agreement with patients’ future MMSE scores. These findings highlight the potential of combining transformer-based speech representations with clinical features to capture subtle markers of cognitive decline over time.

A key contribution of this work is the identification of deep contextual speech representations as important predictors of future cognitive severity. As shown in Figures 5–6, speech features demonstrated a meaningful predictive contribution, with SBERT principal component 1 ranking as the third most important feature in the model (mean feature importance = +0.018). This finding suggests that transformer-based representations of semantic coherence within longitudinal speech transcripts capture clinically relevant information beyond traditional linguistic measures. Specifically, these contextual embeddings may encode subtle changes in discourse organization and coherence associated with cognitive decline. Additionally, baseline diagnosis and the Blessed Dementia Rating Scale ranked among the next most important features after entry MMSE (Figure 6), underscoring that clinical and diagnostic history remain the model’s dominant contributors. The Blessed score specifically captures functional impairment in daily activities among patients with dementia (Blessed et al. 1968), and its high feature importance supports the relationship between functional deterioration and measurable cognitive decline. Together, these findings demonstrate that both speech-derived representations and established clinical assessments provide complementary signals for predicting future cognitive trajectories.

**Fig. 5.**
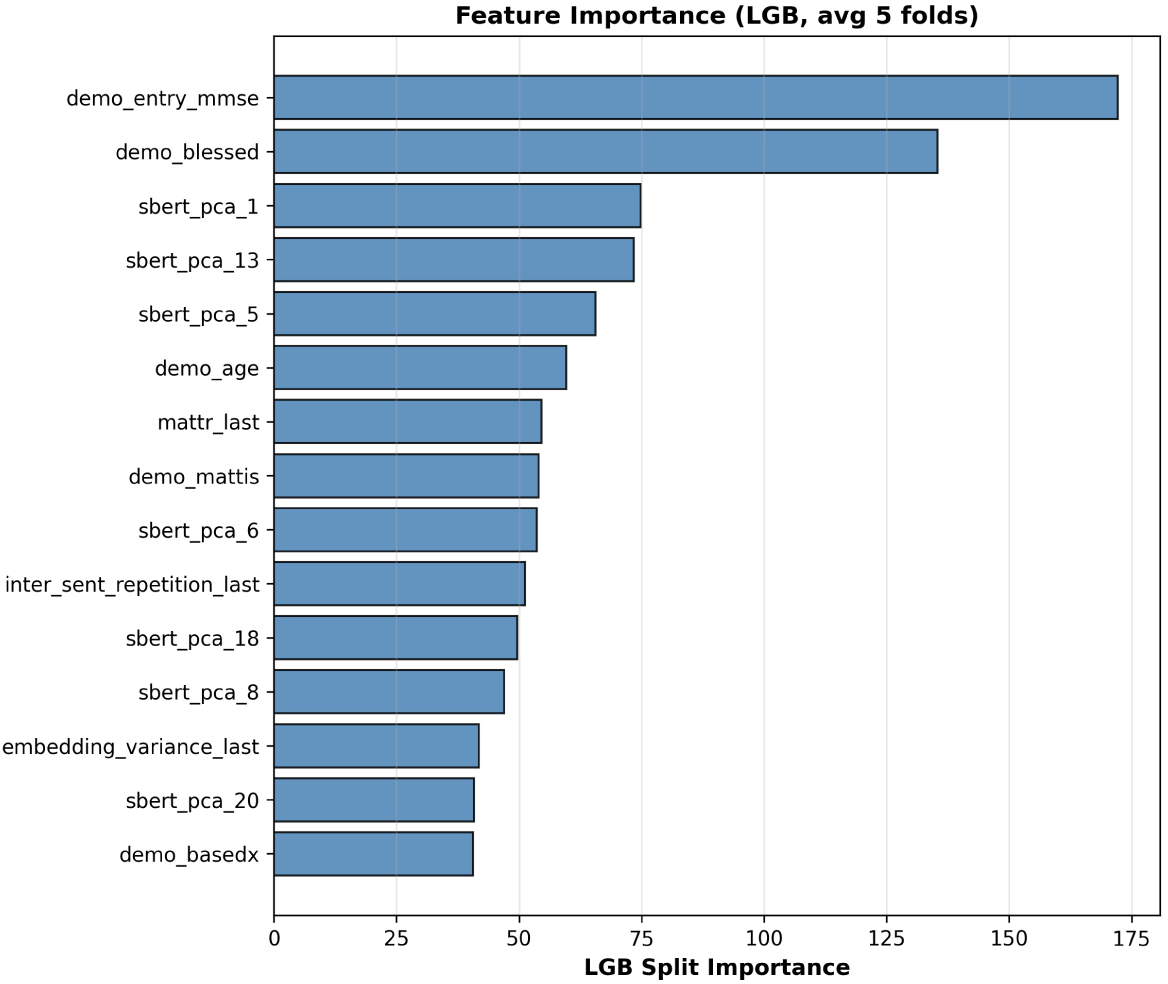
Shows which features the trained LightGBM model relies on most when predicting future MMSE scores, based on the model’s built-in split-based importance score. Bars indicate how often each of the 15 top-ranked features was used to split decision trees, averaged across the 5 cross-validation folds, and include clinical variables such as entry MMSE and the Blessed Dementia Rating Scale alongside components of the SBERT speech embeddings reduced via PCA (sbert_pca_*). Entry MMSE and the Blessed score clearly dominate, while individual speech features each contribute a smaller, fairly even share.

**Fig. 6.**
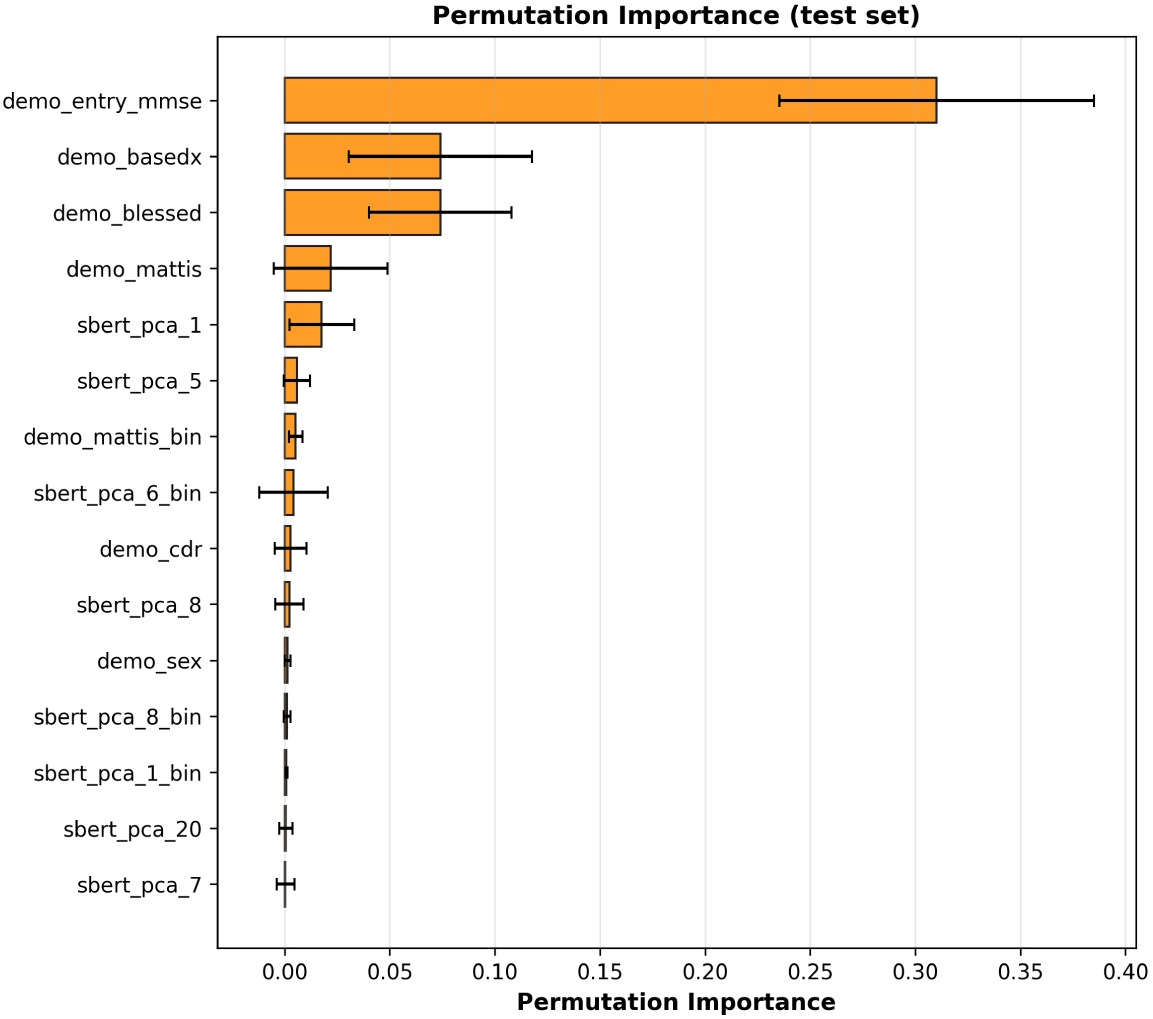
Shows the same features ranked instead by permutation importance, a model-agnostic check that is less biased toward high-cardinality features than the split-based measure in Figure 5. Bars indicate the average drop in out-of-fold *R*^2^ when a feature’s values are randomly shuffled (*̄I_j_* ± σ*_j_*, Eq. 16), with error bars reflecting variability across the 5 folds. Entry MMSE causes by far the largest and most consistent drop in performance when removed, followed by baseline diagnosis and the Blessed score, while speech-derived features contribute smaller but still meaningful importance.

The ablation analysis (Section 3.2) further supported this complementary role. While clinical and demographic features provided the strongest standalone predictive signal (*R*^2^ = 0.716), incorporating SBERT embeddings improved performance beyond clinical features alone, with the combined Clinical + SBERT subset achieving the best feature-subset ablation performance (*R*^2^ = 0.759). This suggests that transformer-based speech representations capture additional information relevant to cognitive function, and that the model’s performance is not driven by clinical variables alone.

### 4.2 Comparison with Prior Work

Prior studies on speech-based dementia detection have shown that transformer-derived speech representations achieve better classification performance than traditional, feature-based approaches. Yang et al. (2022) found in their review of various deep learning approaches that transformer models consistently out- performed traditional machine learning methods for speech-based Alzheimer’s disease classification. Another study conducted by Balagopalan et al. (2020) confirmed this finding, concluding that BERT transformer embeddings consistently outperformed bag-of-words vectorization in dementia detection. Because our study showed that SBERT-derived embeddings had high feature importance relative to handcrafted linguistic features, this pattern is consistent with our findings. However, our framework extends beyond a binary classification task for dementia detection; the high model performance observed with a regression task for predicting future MMSE scores shows that transformer embeddings are valuable for longitudinal severity prediction as well.

Qiao et al. (2021), in a similar fashion to our concatenation of handcrafted discourse features with SBERT-PCA embeddings, tested the accuracy of a hybrid framework combining handcrafted linguistic features with pre-trained transformer embeddings and found an improved dementia detection accuracy compared to using handcrafted features alone. The results of our study extend their hybrid strategy from single-visit classification to multi-visit longitudinal regression on future MMSE scores. Additionally, our framework incorporates clinical and demographic variables, which had the dominant predictive importance; they were the most meaningful in predicting patient cognitive decline. These variables were omitted from the speech-only framework proposed by Qiao et al. (2021).

A study conducted by Balagopalan et al. (2021) compared the performance of pre-trained language foundation models and handcrafted linguistic feature approaches for Alzheimer’s disease detection, finding superior performance for the former. Our feature-importance analysis is consistent with the fact that transformer-derived embeddings hold a meaningful degree of predictive importance; however, our framework combines both handcrafted linguistic feature approaches and transformer-embedding approaches, rather than contrasting them.

Prior work has also included longitudinal analysis of changes in speech biomarkers. Studies by Forbes-McKay et al. (2013) and Gkoumas et al. (2024) both showed that speech biomarkers and linguistic features change significantly across multiple patient visits. However, these studies focused only on observing change, rather than predicting a future state. Our work then constitutes a significant extension: rather than simply noting changes in speech biomarkers, we used linguistic changes over time, together with transformer-derived semantic representations and clinical and demographic variables, to predict a patient’s future MMSE score. All in all, our proposed framework offers a unique angle in the use of speech biomarkers to predict future cognitive states that no single prior study has done: the framework combines transformer-derived semantic representations utilized in cross-sectional dementia detection studies (Yang et al. 2022; Qiao et al. 2021; Balagopalan et al. 2020, 2021) with longitudinal observation explored in previous studies (Forbes-McKay et al. 2013; Gkoumas et al. 2024) for a novel task unexplored directly by either area of work. Rather than simple classification or tracking of change, our task involves accurately predicting future cognitive severity in patients with dementia.

### 4.3 Limitations

This study presents a longitudinal dataset combined with a machine learning framework; however, some minor limitations remain. Firstly, the DementiaBank Pitt corpus dataset used in this study was small; it contained information from only 126 patients, yielding just 173 prediction samples. Importantly, data were limited for patients with severe cognitive impairment (i.e., MMSE *<* 10). As a result, there was a mild overprediction bias for severely impaired patients. The model’s average error (MAE = 2.02, RMSE = 2.75) is also comparable in magnitude to previously reported inter-rater scoring variability on the MMSE itself (Molloy et al. 1991), suggesting the residual error is not clearly distinguishable from the instrument’s own measurement noise. Additionally, our dataset did not include any neuroimaging data, biomarker data, medication history, or other pharmacological data, all of which are known to have independent predictive weight in cognitive decline. As a result, our predictors — though rich within the speech and clinical domains — did not extend to these aforementioned information sources. Adding these information sources, along with benchmarking against simpler baseline models (e.g., linear regression on clinical features alone), provides a natural extension for future modeling.

Finally, our framework was evaluated on a single cohort and was not externally validated on an independent dataset. This is a common constraint in longitudinal speech-based dementia research: a publicly accessible dataset offering longitudinal speech recordings or transcripts, repeated MMSE measurements, and a comparable prediction setting to the one used here was not identified. As such datasets become available, external validation of this framework across additional longitudinal cohorts represents an important direction for future work.

## 5 Conclusion

In conclusion, our framework, which combines transformer-derived semantic embeddings with handcrafted linguistic features, clinical and demographic variables, and temporal characteristics with a gradient-boosted machine learning model, holds strong predictive performance for future MMSE score forecasting. Model results and feature importance outcomes are stable across cross-validation folds. The model shows minimal evidence of overfitting. Based on our feature-importance analysis, clinical features, such as the initial patient MMSE score, are the most influential predictors of cognitive severity progression (Xie et al. 2012). Speech features, such as contextual transformer-derived semantic embeddings, provide a meaningful predictive value; therefore, our results support the view that speech is a promising longitudinal digital biomarker of cognitive decline. An ablation study (Section 3.2) supports this conclusion: the model does not rely on a single feature source, and speech-derived representations provided predictive information complementary to, rather than a substitute for, clinical history. Our contribution addresses the current gap in the literature across related lines of work by moving beyond cross-sectional dementia detection toward predicting future cognitive decline using transformer-derived speech representations and longitudinal patient history. As these findings are drawn from a single cohort, external validation on independent longitudinal datasets will be an important next step toward establishing the generalizability of this framework.

Broader implications of our work include that, because speech collection may be less burdensome for both clinicians and dementia patients than formal cognitive assessments, this framework could help reduce clinician workload, enabling more frequent monitoring. The success of our work in developing an accurate model to predict future cognitive severity progression suggests that using our framework may support better-timed treatment adjustments and improved planning in patient care. Our model can detect small changes in patient MMSE scores between visits, potentially enabling subtle changes in cognitive severity progression to be captured before a patient reaches a clinical threshold for cognitive impairment.

For future extensions of this study, we aim to incorporate larger, more balanced longitudinal datasets that include more samples of patients with severe impairment. Additionally, rather than incorporating a gradient-boosted machine learning model, we aim to implement temporal deep learning architectures, such as LSTMs (Hochreiter and Schmidhuber 1997) and Transformers (Vaswani et al. 2017), to model visit sequences directly. Further, we look to incorporate neuroimaging, biomarker, and pharmacological data into our dataset, as previously discussed. As suitable longitudinal cohorts become publicly available, we also aim to externally validate the trained model on an independent dataset of different patients and differently recorded speech, to assess its ability to generalize. Finally, we look to modify our inclusion of transformer-derived semantic embeddings; rather than deriving these embeddings from a general Sentence-BERT transformer model, incorporating embeddings derived from language models fine-tuned on clinical tasks provides another extension of what we have done in this study.

## Data Availability Statement

The dataset used in this study is available through DementiaBank under controlled access procedures. Due to data-use restrictions, raw speech transcripts and other patient-level records cannot be publicly redistributed; researchers may request access directly through the DementiaBank repository. Source code and analysis pipelines will be made available to facilitate reproducibility.

## Data Availability

The data used in this study were obtained from the DementiaBank Pitt Corpus, which is available to qualified researchers through an access request process. Due to data use restrictions and participant privacy protections, the dataset cannot be publicly shared by the authors. Information regarding access to the dataset is available through DementiaBank. All derived results and analyses supporting the findings of this study are available from the corresponding author upon reasonable request.

https://dementia.talkbank.org/

https://github.com/ddevatha1/dementia-mmse-prediction

## Acknowledgements

Generative AI tools were used during manuscript preparation to assist with drafting portions of the abstract and generating figures. All AI-assisted content was reviewed and verified by the authors, who take full responsibility for the accuracy, integrity, and final content of the manuscript.

The authors acknowledge the DementiaBank Pitt Corpus for providing access to the data used in this study.

